# The Co-production of the Roots Framework: A Reflective Framework for Mapping the Implementation Journey of Trauma-informed Care

**DOI:** 10.1101/2022.04.13.22273691

**Authors:** Steven Anthony Thirkle, Angela Kennedy, Petia Sice, Paras Patel

## Abstract

**Background:** The trauma-informed care programme at the Tees, Esk and Wear Valleys Foundation NHS Trust identified a need to evaluate the ongoing service-wide trauma-informed care implementation effort. An absence of staff, service user and system-related outcomes specific to trauma-informed care presented barriers to monitoring the adoption of trauma-informed approaches and progress over time across the Tees, Esk and Wear Valleys Foundation NHS Trust. This paper describes the co-production of a new self-assessment tool, Roots, a discussion-based framework that facilitates learning and improvement by reflecting on positive or negative examples of trauma-informed services.

**Methods:** Using secondary data obtained from an affiliated national trauma summit and instruments found in literature, domains and items were co-produced with the help of trauma-informed care leads, NHS staff and service users. The research design consisted of community-based co-production methods such as surveys, focus groups, and expert consultations.

**Results:** Adopting trauma-informed care requires enthusiasm and commitment from all members of the organisation. Services must adapt to meet the dynamic needs of staff and service users to ensure they remain trauma-informed; this must be done as a community.

**Conclusions:** Following an extensive co-production process, the Roots framework was published open-access and accompanied by a user manual. Roots can provide both qualitative and quantitative insights on trauma-informed care implementation by provoking the sharing of experience across services.

## 1. Introduction

Trauma-informed care has seen various implementation efforts; many have seen local success; others have failed to provide sufficient evidence (1–4). Trauma-informed care is an evolving organic system model (5,6). It requires a paradigm shift in thinking for service providers to deliver care that is rooted in the understanding of the widespread prevalence and effects of trauma on people (7). An informed conceptualisation of trauma is threefold: the trauma *event*, acknowledging that traumatisation can occur when psychological/social integrity is threatened; how the event is *experienced*; and the *effects* of the event (7). This shift in thinking and conceptualisation of trauma constitutes trauma-informed care. All members of the organisation must embrace the constituents of trauma-informed care for the system model to be of significant benefit; it must not be imposed upon individuals but rather emerge from individuals who experience the organisation (8,9). For systemic adoption, many organisations require concrete evidence of the benefits that trauma-informed care provides (10,11). These benefits are numerous; however, they are often not quantifiable. This does not serve the implementation of trauma-informed care well, as service providers are requesting effective techniques for implementing the necessary changes and specific examples of what it means in practice (10). Establishing the need to develop metrics that can be used in qualitative and quantitative ways to demonstrate the effective implementation of trauma-informed care and realise the benefits that this systemic and individualistic change can provide (11). Calling for a development process bespoke to the organisation, as trauma-informed care is an emergent paradigm (3,8).

Organisational culture change demands individuals to follow suit and to sometimes abandon personal principles (12–14). A frictionless change requires the consideration of affect, sub-groups, personal and existing organisational values, and the quantity and quality of support from leadership (8,9). Factoring in complexity concepts such as emergence, self-organisation, and the sensitivity to initial conditions can assist in the understanding of human systems and utilising previous developments can save time and help co-construct a bigger picture of what it means to be trauma-informed (15,16). This article provides basic information on the Roots framework so that future work can benefit; it uses a similar reporting structure provided by Jung et al. (2009); as a result, readers are given a topographical view of the framework (9).

## 2. Background

At the Tees, Esk and Wear Valleys (TEWV) Foundation NHS Trust, Dr Angela Kennedy, with the help of other key figures, established the trauma-informed care programme to implement trauma-informed care into services. Large-scale training and service-change efforts were proving successful. However, an efficient implementation and evaluation method was seen to be missing. Research and development efforts were identified as being necessary to investigate potential solutions. The project that emerged had one aim: To co-produce an integrative framework for data collection, analysis, and interpretation. The objectives to reach this aim were threefold. The first was to identify a relevant body of knowledge and investigate similar approaches that have sought to evaluate trauma-informed care. The second was to co-produce an evaluation framework that is bespoke to the United Kingdom. The third was to produce documentation for practical use. Three fields from the literature helped construct the narrative. These were organisational culture change, complexity theory, and trauma-informed care. The organisational culture change literature is highly applicable to trauma-informed care (8,9). Trauma-informed care can be viewed as a culture and the change required is a cultural one. Complexity theory can offer an informative view of culture - recognising that culture is a complex phenomenon that emerges in the interactions between actors is conducive to successful change (15,16,18). Within the trauma-informed care literature, previous frameworks were identified; these were systematically evaluated (1–3,19). The Roots tool emerged from a co-production approach to development with staff and service-users being consulted at each stage.

### 2.1. The Implementation of Trauma-Informed Care

Trauma-informed care is socially constructed in the environment (9). It is manifested and brought to life by the inhabitants of the service. Significant system change is required for the implementation of trauma-informed care. This change marks a shift in thinking from *what is wrong with you* to *what happened to you*. This is an essential step to begin the narrative of care (7). All facets of the organisation must engage in this transformation for trauma-informed care to recognise success. Adaptations to the physical environment, raising awareness, and training staff are relatively minor steps in comparison to the tidal-wave requirements of change on systems and care processes (20).

### 2.2. Roots – A Discussion-Based Framework That Facilitates Learning and Improvement by Reflecting on Positive or Negative Examples of Trauma-informed Services

Roots is a developmental framework that uses insights from organisational culture change, human behaviour, complexity theory, and trauma-informed care evaluation. Roots was developed in the United Kingdom and was released in April 2021. There are currently two versions available. One for staff and another for service users. Both versions are identical with only changes made to terminology for accessibility purposes. The definition or conceptual model of Roots is as follows: for an organisation to be trauma-informed, it needs to apply trauma-informed principles and culture in practice. Adopting a systems-wide value model requires enthusiasm and commitment from all members of the organisation. As organisations and individuals within the organisation change, the service must adapt to meet the dynamic needs of staff and service users to ensure they remain trauma-informed (9). Communication and clarification of these values, across teams, departments, buildings, or trusts will assist in providing individuals with the self-knowledge that is often missing, i.e., *how are we adhering to the principles of trauma-informed care that we, as a group, have selected as being relevant right now?* and *how are we able to improve in the areas that we are not doing so well in?* The intended purpose of Roots is to map the implementation journey of trauma-informed care. The tool is for staff, service users, and teams to think about what might make up trauma-informed care in their areas. It takes the form of a word document that is completed by the facilitator on the discussion of each item. Roots is comprised of 54 items answered quantitatively using a RAG (red, amber, and green) rating, and qualitatively by prompting for reasons of applicability and examples in practice. These items are shared among seven domains: Safety (11 items), Language (8 items), Social (7 items), Trauma-specific Interventions (7 items), Empowerment (7 items), Whole System (6 items), and Compassionate Leadership (8 items). The set of practice points for reflection by the group challenges thinking and enables discussion. The *applicable to service* column is asking for the reason why this item needs to be applied in service to facilitate trauma-informed care. However, each item is indicative and may not be applicable in every setting. The *implementation* column is the RAG rating which asks the user how trauma-informed they believe their service is with regards to the item in question. The *example* column asks for examples as to why the service may or may not be delivering trauma-informed care. Reflecting on each practice point can stimulate positive or negative examples and provide meaningful information. The act of assigning a colour can allow the individual or service to reflect on their current standing with trauma-informed service delivery. This can also prompt and motivate individuals and services to improve delivery. Providing examples can be useful for clarity and comparison. The level of measurement used in Roots is ordinal.

Items were generated by using a combination of surveys, focus groups, expert meetings, and secondary data. The domains within Roots were obtained from the National Trauma-informed Care Community of Action’s implementation report titled: *Creating a Narrative for Trauma-Informed Service Transformation* which emerged from a summit of clinicians, managers, leaders, people with lived experience, researchers, and other interested parties on Thursday 28th March 2019 (21). An investigation into the literature uncovered previous approaches. These instruments and frameworks were studied, and domains were taken from four prominent frameworks which were then compared at a trauma leads meeting with the domains that emerged from the national trauma-informed care summit. At the same meeting, trauma leads were provided with definitions on the summit domains and asked to provide a set of standardised questions (items). These items were then shared with psychologists to translate for staff accessibility purposes. Further translation was required for the service-user version and so the staff version was shared with an expert by experience group. Both staff and service-user versions of the framework were then issued to wider audiences at the NHS trust using surveys to further articulate the forms. Trauma leads meetings were used to discuss the results and changes were decided on. Focus groups were then arranged to pilot test the framework. These were conducted with staff; trauma leads and their staff members in their respective services, and a small group of service-users. Prior to the focus groups, the service-user framework was modified to reflect a third-person perspective rather than what was seen to be a confusing first-person one; this was the only significant modification made to the framework since the initial translation. Items were not reduced or modified further as they were voted as being essential to remain in the framework. Due to resource restrictions, it was not possible to conduct a follow-up assessment on the psychometric properties of an instrument (reliability and validity). However, focus group data would suggest strong face validity, acceptability, and feasibility. The framework does have the potential to be susceptible to bias if the facilitator is part of the team or has an agenda. This can be mitigated by using external facilitators. There are no norms attached to the Roots tool. However, these can be created locally if group comparisons are desired. Roots has no formal method of calibration. Informal methods can be set by users of the tool to ascertain accuracy. There is currently no data recorded for further tests of reliability such as internal consistency or reproducibility (test-retest and inter-observer). The content, criterion, predictive, convergent, discriminant, cross-cultural, and dimensional validity of Roots remains untested. Roots is designed to be sensitive and open to change, the tool must reflect the needs of the individuals it serves.

The Roots tool was co-developed, and pilot tested in mental health care settings. These services included the community, prison services, adult wards, and CAMHS (Child and Adolescent Mental Health Services). However, Roots has the potential to be deployed in any setting, with changes made to the language and the exercise.

## 3. Statement of the Problem

The Tees, Esk and Wear Valleys NHS Foundation Trust has been implementing trauma-informed care for many years. Early implementation efforts took the form of care pathways. In 2009, the trauma-informed pathway was designated as the first clinical link pathway. This pathway differed from clinical routes in that it was relevant regardless of the diagnosis of trauma. Instead of encouraging talents that the workforce cannot supply, the trauma-informed pathway encourages staff to use their skills in trauma-informed ways. This empowered staff when they realised that this meant they could offer something critical to service users. A business case was developed by the trauma-informed care lead, Dr Angela Kennedy, for a formally funded project to embed trauma-informed care into services. A goal of the project was to integrate trauma-informed care into policies, programmes, and local systems and contribute to the evidence-base for trauma-informed care. The trauma-informed care programme at the Tees, Esk, and Wear Valleys NHS Foundation Trust realised the need for an evaluation and implementation framework to make progress with this trauma-informed system change. Checklists have been created and measurements have been conducted previously regarding trauma-informed change, but never in the United Kingdom. The NHS is a unique system in that it represents a large number of individuals spread out over many localities. The TEWV NHS Foundation Trust is one of the largest specialist mental health and learning disabilities trusts in the country, with an annual income of £380 million and a workforce of over 6700 staff operating from around 100 sites in Durham, Teeside, North Yorkshire, and York and Selby. TEWV NHS provide a range of inpatient and community services to 2 million people living in County Durham, the Tees Valley, Scarborough, Whitby, Ryedale, Harrogate, Hambleton and Richmondshire. TEWV NHS services are spread out over a wide geographical area of around 3600 square miles, inclusive of coastal, rural, and industrial areas (22). The geographical nature of the TEWV NHS Foundation Trust is conducive to complexity. All NHS trusts are subject to stringent ethical procedures and research involving staff and service-users undergo strict ethical clearance procedures. An implementation and evaluation framework unique to these particular circumstances that was able to navigate strict ethical boundaries was essential.

## 4. Methods Used During the Development of the Roots Framework

The collaborative development of the Roots framework was a five-step process. Firstly, a literature review was undertaken to investigate similar approaches. These other approaches were evaluated using an existing culture instrument review framework (17), and were then compared with the domains that emerged from the national trauma summit at a trauma leads meeting. Items were generated at the same meeting and were taken to surveys with staff and service users to confirm articulation. Focus groups were held with one service user group and five different staff services. These were used to further articulate and pilot test the framework in practice. The process can be seen in Figure 1.

**Fig. 1.**
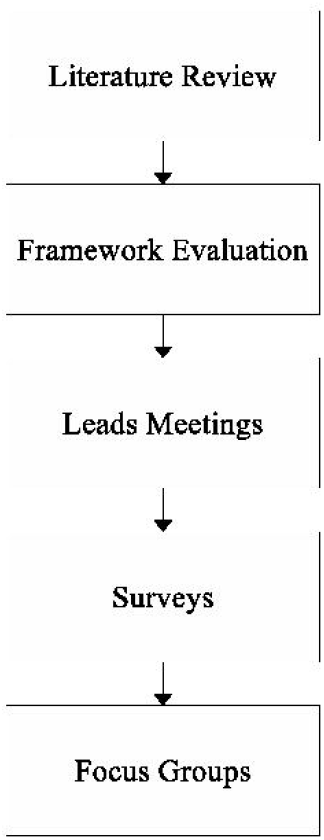
Methods used during the co-production process of Roots

The design of the Roots framework stemmed from the ongoing work of the trauma-informed care programme. This meant that the research design evolved alongside programme implementation. Monthly meetings with trauma leads contributed significantly to the research design. The trauma leads are select individuals who represent trauma-informed care in their respective services. They were instrumental in the facilitation of various developmental facets, including access to various staff and service user groups. Approximately twelve trauma leads would usually be present at each meeting. The first phase of this study was an evaluation of other similar instruments and frameworks (9). The literature contributed to expectations and other considerations for trauma-informed service evaluation; including the use of principles or domains for which both implementation and evaluation can revolve. Meanwhile, the trauma-informed programme was a joint-organiser of a national trauma summit titled: *Creating a Narrative for Trauma-informed Service Transformation* (21). The goal of this summit was to respond to calls from people with lived experience of trauma in services and the challenge of trauma being included in the new NHS 10-year plan. It was arranged as a forum to share ideas for good practice and an opportunity to network with others who are motivated towards similar goals. This was held on Thursday 28^th^ March and brought together clinicians, managers, leaders, people with lived experience, researchers, and many others. Attendees were asked to think of a specific positive example that they have experienced, witnessed, or been involved in. The aim of this was to tap into individual wisdom and creativity and to explore the multiple dimensions of what it means to be trauma-informed and share concrete examples that can be replicated by others. A report was drafted titled: *Developing Real-World System Capability in Trauma-informed Care: Learning from Good Practice* (21). In this report, seven domains were observed as having emerged from the summit, these were: 1) Safety, 2) Human Experience Language, 3) Empowerment, 4) Healing Interventions, 5) Responsive System Design, 6) Compassionate and Transformational Leadership, and 7) Relational Reparation (21). These domains were significant to the Roots framework as they had been developed with individuals that have experienced UK health systems. The domains were used but their wording was amended to better reflect local services. The revised domains were: 1) Safety, 2) Language, 3) Social, 4) Trauma-specific Interventions, 5) Empowerment, 6) Whole System, and 7) Compassionate Leadership.

## 5. The Co-Production of Domains and Items

At a trauma leads meeting, the seven domains were judged against the domains taken from the literature. Four instruments were selected for this process, the Attitudes Related to Trauma-Informed Care (ARTIC) (1), the Creating Cultures of Trauma-informed Care (CCTIC) (4), the TICOMETER (2), and the Trauma-informed Practice (TIP) Scales (19). These were identified as being relevant and empirically tested. During the meeting, the trauma leads worked through a slideshow that presented the domains from the literature alongside the domains from the summit and were asked to determine encapsulation. Encapsulation was confirmed if one or more of the summit domains captured the meaning used in the language of the domains from the literature. Table 1 presents the ARTIC comparison, Table 2 presents the CCTIC comparison, Table 3 presents the TICOMETER comparison, and Table 4 presents the TIP Scales comparison. The left column represents the domains from the instruments found in the literature, and the right column represents which domain from the summit that trauma leads felt captured the meaning of the corresponding domain from the literature. This process was held to determine if the summit domains were appropriate for use. The trauma leads all agreed through consensus that all domains from the four instruments were captured by one or more of the domains from the summit. However, trauma leads felt as though staff wellbeing should represent the eighth domain.

**Table 1.**
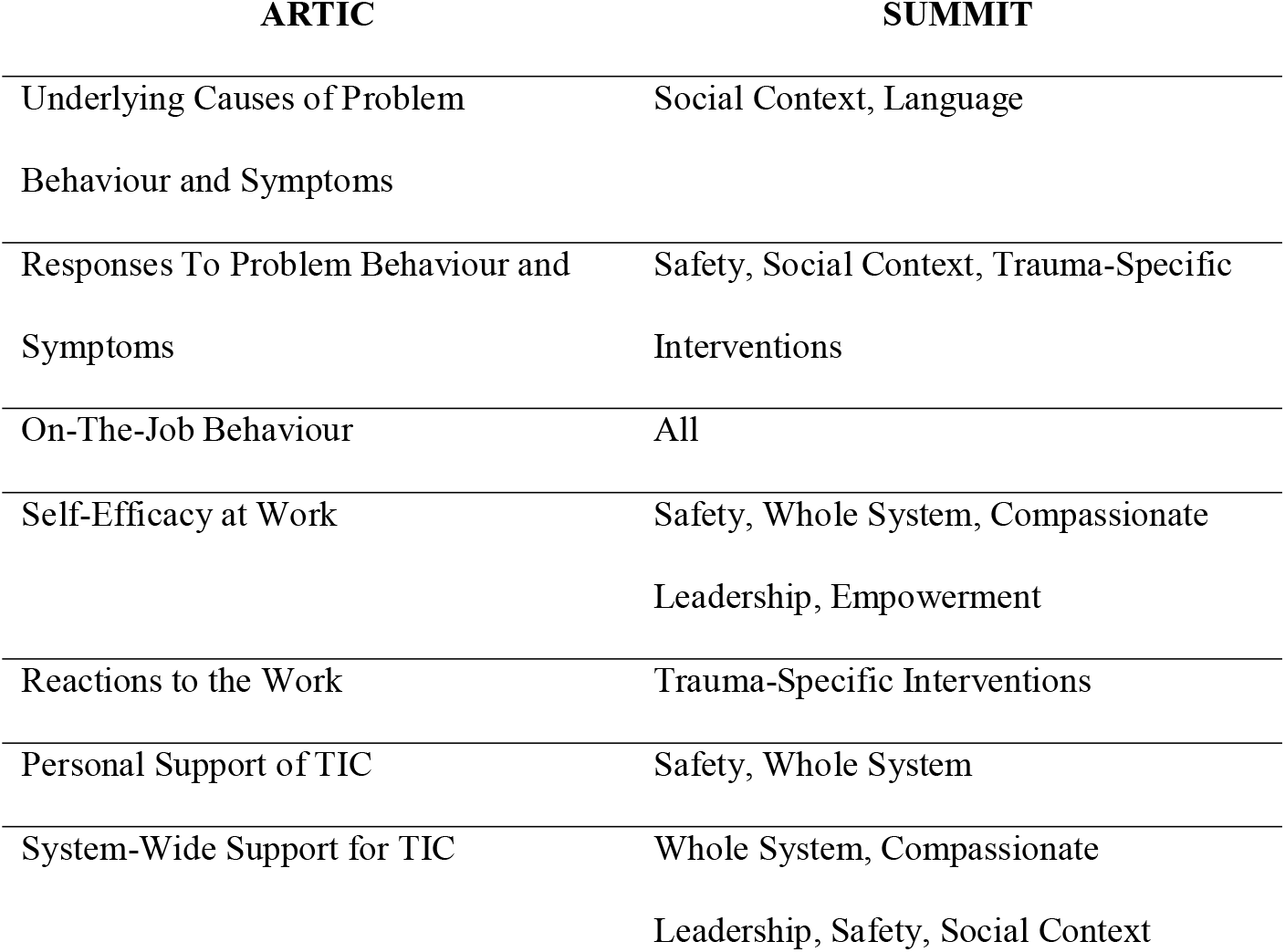
ARTIC Comparison

**Table 2.**
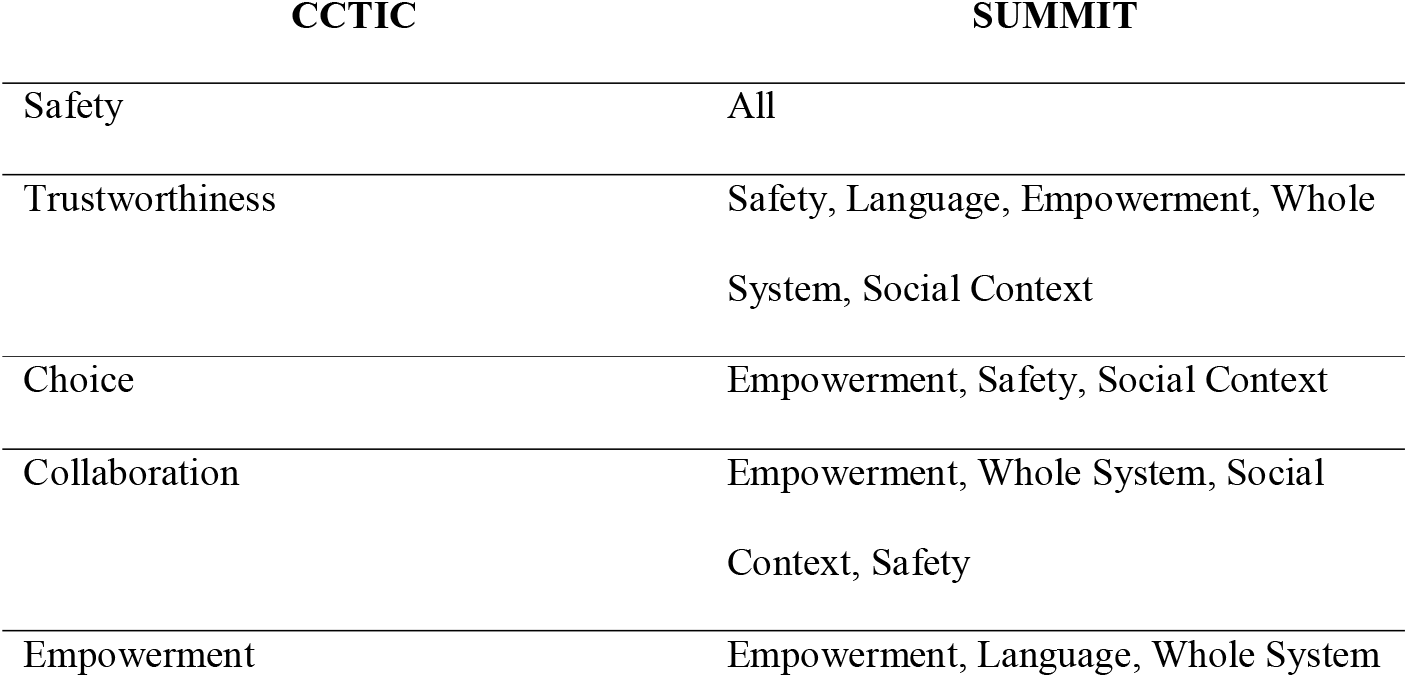
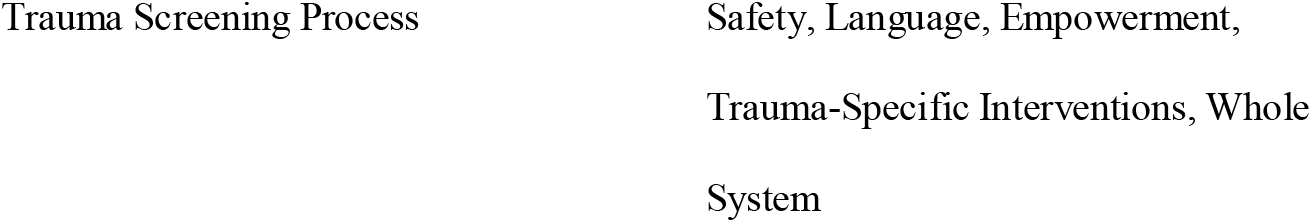
CCTIC Comparison

| CCTIC | SUMMIT |
| --- | --- |
| Safety | All |
| Trustworthiness | Safety, Language, Empowerment, Whole System, Social Context |
| Choice | Empowerment, Safety, Social Context |
| Collaboration | Empowerment, Whole System, Social Context, Safety |
| Empowerment | Empowerment, Language, Whole System |

**Table 3.**
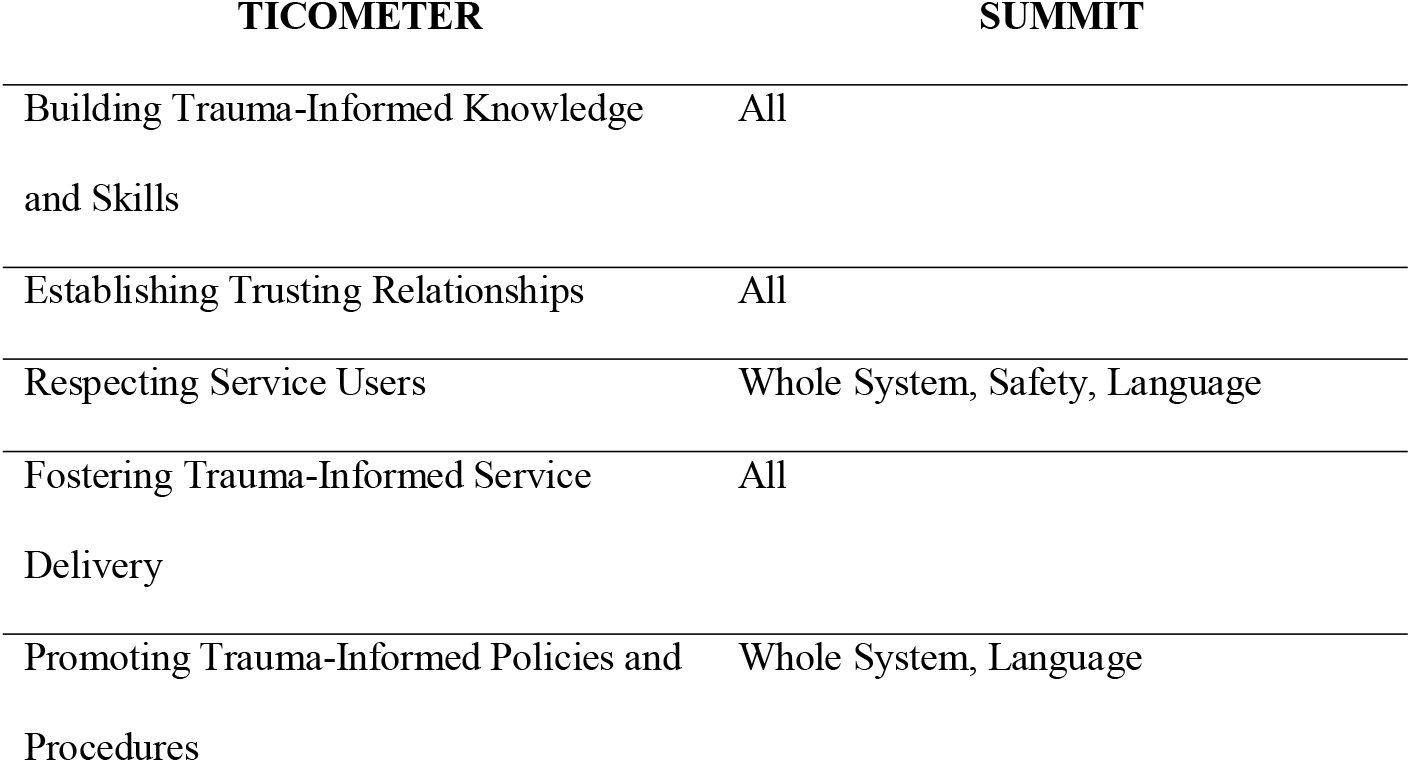
TICOMETER Comparison

**Table 4.**
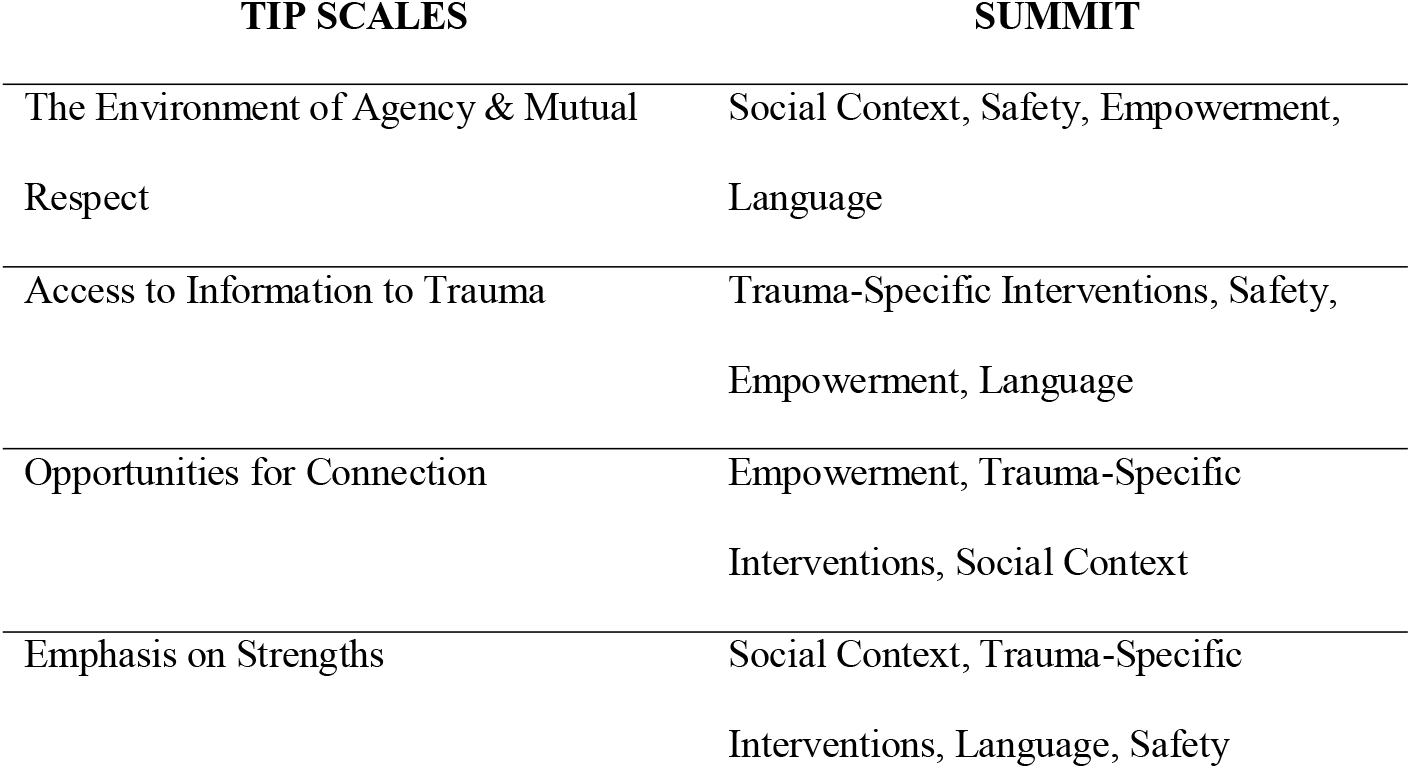
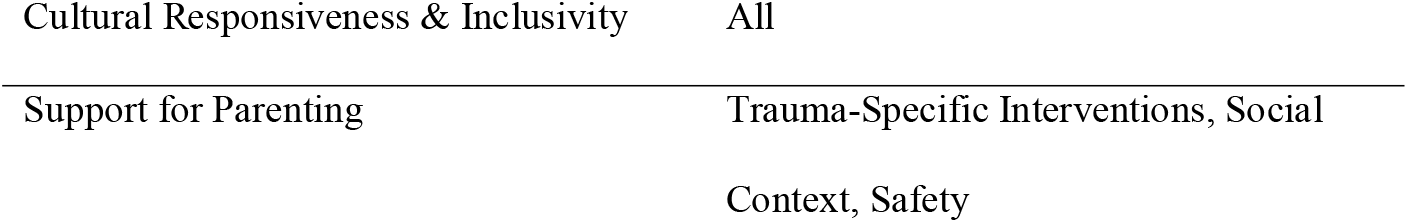
TIP Scales Comparison

At the same meeting, trauma leads were asked to develop up to 10 items per domain. Trauma leads worked through a slideshow that presented definitions of each domain. Four items were generated for the domain safety. Six items were generated for the domain language. Four items were generated for the domain social. Six items were generated for the domain Trauma-specific Interventions. Eleven items were generated for the domain empowerment. Seven items were generated for the domain Whole System. Eight items were generated for the domain compassionate leadership. Four items were generated for the domain staff wellbeing.

The results of this meeting were shared with the trauma-informed care programme team. The items underwent translation to accommodate common language used by staff for purposes of familiarity and accessibility. This discussion contributed to the removal of some items and the addition of others. The requested domain, staff wellbeing, was removed as it was voted unnecessary in respect of it being represented across the other domains. As two forms were needed, one for staff and another for service users, the translated items were then sent to an experts by experience group working from the Recovery College in Durham, United Kingdom. The experts by experience group worked through the items and translated them for service user accessibility.

Both forms, existing only as items, were then placed into two separate SurveyMonkey surveys. Convenience, criterion, and snowball sampling methods were used for the recruitment of participants. Staff and service-user samples were recruited through trauma leads who distributed information on the study. When participants registered interest, the trauma leads then distributed further information through a participant information sheet and a web link to the survey. The surveys asked participants to prioritise the items and leave feedback on articulation. The addition of a RAG rating added a quantifiable indicator to the survey. The staff form was distributed to staff members, and the service-user form was distributed to service users. The inclusion criteria were as follows: staff are identified as being available, engaged in the TEWV NHS trust, and chosen or recommended by those aware of the study. Service users are identified as being available, involved in the NHS trust, and selected by staff. The exclusion criteria were as follows: no affiliation with the NHS TEWV Foundation Trust, under the age of 18 or over the age of 65, or at imminent risk of harming themselves or others. Recruitment was difficult as national restrictions were imposed as a result of the Covid-19 pandemic. The results were analysed using Microsoft Excel to determine priority. A trauma leads meeting was held to discuss these results and it was voted that all items should remain in the final framework, mainly due to high applicability. Although the survey results contradicted this for a few items, the presentation of the survey was questioned for confusion.

A quality assurance process took place and involved confirming meaning across items in both staff and service user forms with the research team. The framework was drafted as a table, in which there are four columns: practice point for consideration (item), applicable to service (reason), implementation status (RAG rating), and example (justification). The set of practice points for reflection by the group challenges thinking and enables discussion. The applicable to service column asks why this item needs to be applied in service to facilitate trauma-informed care. However, it is recognised that each item is indicative and may not be applicable in every setting. The implementation column is the RAG rating and questions the user on how trauma-informed they believe their service is with regards to the item in question. The example column asks for examples as to why the service may or may not be delivering trauma-informed care. Reflecting on each practice point can stimulate positive or negative examples and provide meaningful information. The act of assigning a colour can allow the individual or service to reflect on their current standing with trauma-informed service delivery. This can also prompt and motivate individuals and services to improve delivery. Providing examples can be useful for clarity and comparison.

Pilot tests were held with both forms with staff and service user sample groups. Five full-length evaluation exercises were held with staff, and one rudimentary exercise was held with service users. Staff focus groups were evaluating the framework and the exercise, whilst the service user group was evaluating the framework. Service users felt empowered when discussing the items of the framework and agreed that the majority of items were essential. Staff found the exercise helpful, and all services requested the completed framework to begin actioning items.

A user manual was created to facilitate instructions and to contain the Roots framework. This is now published open access on the Future NHS community platform for practitioners and researchers as long as they have an NHS or public health England email address.

## 6. The Roots Framework Learning Model

The Roots framework uses insights from complexity theory. This is recognising that human change is complex because there is rarely one right way of doing something. The service will need to adapt to meet each individual’s needs and remain responsive over time. These complexity principles guide the use of Roots towards change and evaluation: change in individuals or organisations is rarely linear. A reflexive approach that evolves is of benefit. There is not a *one size fits all* interpretation of trauma-informed implementation. Different settings need to define what is needed for them through methods of co-production. Different teams within an organisation, different individuals within teams, and different service users may all display or perceive different strengths concerning the implementation of trauma-informed care. Roots allows for the bringing together of different narratives towards a wider picture. The items emerged from previous examples and are created to be both generic and specific but not exhaustive. New ones can be created as long as they are tangible and observable. Roots needs to be embedded within a learning organisation framework accompanied by an attitude of respect to ensure that progress can be made.

## 7. Study Limitations

In the UK, there are many restrictions placed upon social and healthcare research. Research studies must navigate strict ethical standards. In many cases, these standards can form boundaries for what is possible in research. As trauma-informed care is a relatively new concept in the UK, pioneering research must first take the first steps. These initial studies must take place so that more elaborate studies can follow. As Roots is the first trauma-informed evaluation framework to emerge out of the UK, many aspects of the study were impeded. The Tees, Esk and Wear Valleys NHS Foundation Trust has a workforce of over 6700 staff operating from around 100 sites in Durham, Teesside, North Yorkshire and York and Shelby (22). This study is unable to claim to be fully representative as the samples used were often limited to convenience, and the actual number of participants was low. To achieve a confidence level of 99% for a population size of 6700 with a 5% margin of error, 604 staff members must have been involved in the study. For service users, an ideal sample size with the same parameters would be inclusive of 663 participants. In combination, a repeated study would need to consult at least 1200 participants to be statistically significant. The use of qualitative data helps mitigate this through practicality and transparency. The research team had ultimate control over the end-product, meaning that key stakeholders at both staff and service user levels should have been involved in signing off the product. The service user focus group is a good example of this, there were questions raised over some items, but this was not regarded in the finished version. The development of Roots also took place during the Covid-19 pandemic and data collection began when the UK was put under lockdown – this caused delays to the study as all non-covid related research received suspensions from the NHS. There is also a distinct lack of any form of psychometric testing – delays to research and resource expiration made it impossible to continue working on Roots.

## Further Work

Roots is the United Kingdom’s first foray into the evaluation of trauma-informed services. Much of the precursory work is already done in the United States of America (USA). However, UK healthcare services are delivered very differently than they are in the USA. The development of Roots provides perfect foundations for future work, either using or building on Roots. One of the fundamentals of UK-based development is the practicalities of set-up. Establishing long-term commitment and navigating strict ethical standards are two of the arrangements that must be considered. Acquiring health research authority approval through a completed Integrated Research Application System (IRAS) should be completed early, and applications should be thorough to ensure acceptance. Before use, a thorough psychometric assessment of the tool should be prioritised to ensure reliability and validity. Self-assessment maintenance should be carried out with exceptional regard to the language used. A dynamic self-assessment should be co-produced; the contents of which change with time to ensure consistent reliability with the people they serve.

## Conclusions

The co-production of the Roots framework involved staff and service-users from the Tees, Esk and Wear Valleys Foundation NHS Trust and also utilised secondary data from a national trauma summit titled: *Creating a Narrative for Trauma-informed Service Transformation*. Relevant secondary data, trauma-informed care programme lead meetings, surveys, focus groups, and consultations with staff and service users constituted the research design. The development of Roots was an experimental process and the research evolved alongside the progression of the trauma-informed care programme at the Tees, Esk and Wear Valleys Foundation NHS Trust. Roots is published open-access and is supported by a user manual.

## Data Availability

All data produced in the present study are available upon reasonable request to the authors

## DECLARATIONS

### Ethics approval and consent to participate

The United Kingdom’s Research Ethics Committee of the National Health Service’s Health Research Authority gave ethical approval for this work. Informed consent was obtained from all individuals included in the study where necessary.

### Consent for publication

The authors affirm that human research participants provided informed consent where necessary for publication of data collected.

### Availability of data and material

Not Applicable

### Authors’ Contributions

- CONCEPTION: ST, PS and AK
- METHODOLOGY: ST, PS, AK and PP
- DATA COLLECTION: ST
- INTERPRETATION OR ANALYSIS OF DATA: ST
- PREPARATION OF THE MANUSCRIPT: ST
- REVISION FOR IMPORTANT INTELLECTUAL CONTENT: ST, PS, AK, and PP
- SUPERVISION: PS, AK, and PP

## Acknowledgements

This work was made possible by support from the Tees, Esk and Wear Valleys Foundation NHS Trust and Northumbria University..

